# Apolipoprotein E moderates the association between Non-*APOE* Polygenic Risk Score for Alzheimer’s Disease and Aging on Preclinical Cognitive Function

**DOI:** 10.1101/2023.06.09.23291215

**Authors:** Yuexuan Xu, Zhongxuan Sun, Erin Jonaitis, Yuetiva Deming, Qiongshi Lu, Sterling C. Johnson, Corinne D. Engelman

**Affiliations:** Department of Population Health Science, School of Medicine and Public Health, University of Wisconsin-Madison; Department of Biostatistics and Medical Informatics, School of Medicine and Public Health, University of Wisconsin-Madison; Department of Medicine, School of Medicine and Public Health, University of Wisconsin-Madison; Wisconsin Alzheimer’s Institute, University of Wisconsin-Madison; Wisconsin Alzheimer’s Disease Research Center, University of Wisconsin-Madison

**Keywords:** PRS, *APOE*, aging, interaction, cognition

## Abstract

**INTRODUCTION:** Variation in preclinical cognitive decline suggests additional genetic factors related to Alzheimer’s disease (e.g., a non-*APOE* polygenic risk scores [PRS]) may interact with the *APOE* ε4 allele to influence cognitive decline.

**METHODS:** We tested the PRS×*APOE* ε4×age interaction on preclinical cognition using longitudinal data from the Wisconsin Registry for Alzheimer’s Prevention. All analyses were fitted using a linear mixed-effects model and adjusted for within individual/family correlation among 1,190 individuals.

**RESULTS:** We found statistically significant PRS×*APOE* ε4×age interactions on immediate learning (*P*=0.038), delayed recall (*P*<0.001), and Preclinical Alzheimer’s Cognitive Composite 3 score (*P*=0.026). PRS-related differences in overall and memory-related cognitive domains between people with and without *APOE* ε4 emerge around age 70, with a much stronger adverse PRS effect among *APOE* ε4 carriers. The findings were replicated in a population-based cohort.

**DISCUSSION:** *APOE* ε4 can modify the association between PRS and cognition decline.

**Highlights:**

- *APOE* ε4 can modify the association between PRS and longitudinal cognition decline, with the modifying effects more pronounced when the PRS is constructed using a conservative *P*-threshold (e.g., *P* < 5^e-8^).
- The adverse genetic effect caused by the combined effect of the currently known genetic variants is more detrimental among *APOE* ε4 carriers around age 70.
- Individuals who are *APOE* ε4 carriers with high PRS are the most vulnerable to the harmful effects caused by genetic burden.

## 1. INTRODUCTION

Late-onset Alzheimer’s disease (LOAD) is an age-related neurodegenerative disease with neuropathologic changes decades before overt symptoms. It is critical to have a low-cost, non-invasive test to identify the population at risk during this preclinical phase for clinical trial eligibility and early biomarker screening. To this end, genetic risk factors are a promising tool for risk prediction and stratification in clinical settings. The *APOE* gene is the strongest genetic risk factor for LOAD, with the *APOE* ε4 allele conferring increased risk and the *APOE* ε2 allele conferring reduced risk [1]. Additionally, recent meta-analyses of genome-wide association studies (GWAS) have identified dozens of single-nucleotide polymorphisms (SNPs) outside the *APOE* region that are associated with the risk of LOAD [2–5]. However, these SNPs exhibit miniscule effects on AD risk, and, therefore, the effectiveness and accuracy of prediction using any single genetic variant is limited. Polygenic risk scores (PRSs), on the other hand, sum the effects of multiple independent SNPs, converting the overall genetic burden to a single score. PRSs have been utilized in multiple AD risk prediction models, serving as effective predictors of disease risk, with up to 84% accuracy [6].

The genetic risk of LOAD measured by *APOE* and non-*APOE* PRS is age-related. In a prospective cohort of 1,200 asymptomatic individuals, our group found that a statistically significant effect of *APOE* on beta-amyloid (Aβ) emerged around age 55, on tau emerged at age 65, and on cognition emerged at age 65–70. Similar age-related trajectories were observed for non-*APOE* PRS; however, the effect emerged approximately a decade later than the *APOE* effect (e.g., age 65 for Aβ, age 70 for tau, and age 75–80 for cognition) [7]. These findings are consistent with recent studies using Alzheimer’s Disease Neuroimaging Initiative (ADNI) and UK Biobank (UKBB) data, concluding that both *APOE* and non-*APOE* PRS predicted LOAD risk and presented age-related effects, but the *APOE* effects were stronger among individuals younger than 80 years of age, whereas the non-*APOE* PRS effects were stronger for those older than 80 years of age [8]. Similar findings were also reported in UKBB on the association between *APOE*, non-*APOE* PRS, and longitudinal cognition [9].

In addition to age-related genetic effects, it has been shown that the contribution of non–*APOE* PRS to LOAD also differs according to *APOE* ε4 carrier status; however, such findings are not consistent. In a population-based sample of cognitively unimpaired individuals around age 70, an interaction between *APOE* ε4 and non-*APOE* PRS was observed on Aβ42, where the association between non-*APOE* PRS and Aβ42 was only statistically significant among *APOE* ε4 carriers [10]. Similarly, in the Rotterdam study and two other Amsterdam cohorts, the joint effect of common genetic variants on risk of LOAD was stronger among *APOE* ε4 carriers [11,12]. Conversely, in another population-based sample of individuals aged 70 to 111, Najar et al. reported a modifying effect of *APOE* ε4 on the association between non-*APOE* PRS and dementia, but in the opposite direction, such that non-*APOE* PRS only predicted dementia among *APOE* ε4 non-carriers [13]. Even though a growing body of literature has described evidence that *APOE* can modify the effect of non-*APOE* PRS on various of AD outcomes, some recent studies failed to find statistically significant interactions between *APOE* and non-*APOE* PRS [14–16].

To date, only a few studies have examined whether and how the risk of non-*APOE* PRS depends on both age and *APOE* and how they jointly affect an individual’s liability to develop LOAD. For example, Fulton-Howard et al. employed data from the Alzheimer’s Disease Genetics Consortium and found a statistically significant interaction between age, non-*APOE* PRS (*P* threshold = 1e^-5^), and *APOE* ε4, with younger *APOE* ε4 carriers bearing greater detrimental effect from the non-*APOE* PRS. However, to our knowledge, no studies have examined the interaction between *APOE,* non-*APOE* PRS, and age on preclinical phenotypes among asymptomatic individuals. To address this question, we tested the associations between non-*APOE* PRS x *APOE* x age and longitudinal global and domain-specific cognition among 1,190 middle-aged, initially cognitively unimpaired individuals from the Wisconsin Registry for Alzheimer’s Prevention (WRAP). We also replicated our focal findings in the Health and Retirement Study (HRS).

## 2. METHODS

### 2.1 Study participants

Data were from individuals enrolled in WRAP, an ongoing prospective longitudinal cohort study of >1,500 late-middle-aged adults who were English speakers and cognitively healthy at enrollment. The WRAP sample is enriched for participants with a parental history of AD, increasing the proportion of individuals who will experience LOAD pathology and cognitive decline during the course of the study and enhancing the ability to identify factors that modify LOAD risk. More than 70% of WRAP participants had a parent with either autopsy-confirmed or probable AD as defined by the National Institute of Neurological and Communicative Disorders and Stroke and the Alzheimer’s Disease and Related Disorders Association research criteria. The details of the study design have been described elsewhere [17]. Briefly, the WRAP study began recruiting participants in 2001, with an initial follow-up after four years and subsequent follow-up biennially. Participants were age 40 to 65 years at baseline. At each study visit, WRAP participants were given an extensive battery of neuropsychological tests, along with a physical and health examination. The maximum number of WRAP visits available at the time of analysis, using the May 2021 data freeze, was seven. WRAP participants who were self– reported non-Hispanic white (NHW) and cognitively unimpaired at baseline with complete cognition, genetic, and demographic data were included in the current study (*N* = 1,190). We excluded data from the first WRAP visit because the global and domain-specific cognitive outcomes that we investigated in the current study cannot be computed using the neuropsychological tests administered in the first WRAP visit. All subjects provided signed informed consent before participation. This study was approved by the University of Wisconsin–Madison Institutional Review Board and was conducted in accordance with the Declaration of Helsinki.

### 2.2 Neuropsychometric assessments

WRAP participants completed cognitive assessments at each WRAP visit. In the current study, we assessed the global cognitive performance of WRAP participants using Preclinical Alzheimer’s Cognitive Composite 3 (PACC-3), which is also the primary outcome in this study, based on work by Donohue and colleagues [18]. Specifically, we computed the PACC-3 score by standardizing and averaging performance from three tests: Rey Auditory Verbal Learning (RAVLT; Trials 1–5), Wechsler Memory Scale-Revised (WMS-R) Logical Memory II total score (LMII), and Wechsler Adult Intelligence Scale-Revised (WAIS-R) Digit Symbol Coding, total items completed in 90 seconds [19]. In addition to testing global cognitive performance, we additionally constructed three domain-specific cognitive scores by averaging standardized test scores, as secondary outcomes, which include immediate learning, delayed recall, and executive function [20]. The immediate learning composite score was derived from RAVLT total trials 1–5, the WMS-R Logical Memory I total score (LMI), and the Brief Visuospatial Memory Test-Revised (BVMT-R) immediate recall score. The delayed recall composite score was obtained based on the sum of the RAVLT long-delay free recall score, the WMS-R logical memory delayed recall score, and the BVMT-R delayed recall score. The executive function domain-specific composite score was constructed based on the Trail-Making Test part B (TMT-B) total time to completion, Stroop Neuropsychological Screening Test color-word interference total items completed in 120 seconds, and WAIS-R Digit Symbol Coding. The *z* score for TMT-B was multiplied by –1 before inclusion into the composite so that higher *z* scores would indicate better performance for all tests.

### 2.3 DNA collection, genotyping, and quality control

Details about WRAP genomic data collection and quality control have been described previously [21]. Briefly, the WRAP genetic samples were genotyped using DNA from whole blood samples and the Illumina Infinium Expanded Multi-Ethnic Genotyping Array (MEGA^EX^) at the University of Wisconsin Biotechnology Center. Individuals with inconsistencies between self-reported and genetic sex, and individuals and SNPs with missingness >5% were excluded. Samples from individuals of genetically defined European descent were then imputed using the Michigan Imputation Server and the Haplotype Reference Consortium (HRC) reference panel. Variants with a low imputation quality score (R^2^ < 0.8), with a low minor allele frequency (MAF, MAF < 0.001), or outside of Hardy-Weinberg equilibrium were removed after imputation. PLINK 1.9 was used in quality control[22,23]. A total of 1,198 individuals with 10,499,994 SNPs remained after quality control. The GRCh37 genome build was used.

### 2.4 Polygenic risk score construction and *APOE*

PRSs were computed using the software PLINK 1.9 [22,23]. SNPs were selected using LD clumping (1000 KB, R^2^ = 0.1 and *P* value threshold of 1.0), and the *APOE* region was excluded (hg19 coordinates chr19 from 44412079 to 46412079) [13,24]. The PRS we used in the main analysis is referred to as PRS_Kunkle_sig_ and is constructed by including SNPs that have shown genome-wide significant (*P* < 5e^-8^, excluding *APOE*) association with AD after combined meta-analyses in the most recent diagnosed case-control GWAS by Kunkle et al. [2]. Additional PRSs were constructed and tested in the sensitivity analyses. First, we created a PRS_Kunkle_1e-5_ by including variants that surpassed the *P* value threshold *P* < 1e^-5^, which is the only *P* value threshold for PRS construction investigated by Fulton-Howard et al. [25]. Then we constructed another set of PRSs which include variants that surpassed *P* < 1e^-3^, *P* < 1e^-1^, and *P* < 1 based on the stage one summary statistics of Kunkle et al.’s GWAS meta-analysis, referred to as PRS_Kunkle_1e-3_, PRS_Kunkle_1e-1_, and PRS_Kunkle_1_, respectively. We created another PRS_deroja_sig_ based on SNPs (excluding *APOE*) that have shown genome-wide significant associations with AD in a large genetic association study by merging all available case-control datasets and by-proxy (e.g., proxy phenotypes using parental history of disease) study results [4]. Unlike traditional PRS, this PRS_deroja_sig_ was constructed using a hybrid way by including genome-wide significant variants identified in [4] and effect size taken from the previous International Genomics of Alzheimer’s Project (IGAP)’s studies. The PRS_deroja_sig_ was also validated for the first time in the clinical sample [4,13]. All PRSs were generated by multiplying the number of each effect allele for each variant by its respective weight (natural log odds ratio) and then summing across all variants. All PRSs were standardized with a mean of 0 and standard deviation of 1 and were used as continuous variables in all analyses for ease of comparison. A higher PRS indicates a higher genetic risk for LOAD. *APOE* genotype was first divided into six sub-genotypes (ε2/ε2, ε2/ε3, ε3/ε3, ε2/ε4, ε3/ε4, and ε4/ε4) based on rs7412 and rs429358 and then combined into two groups that include individuals who are *APOE* ε4 carriers (ε2/ε4, ε3/ε4, ε4/ε4) and non-carriers (ε2/ε2, ε2/ε3, ε3/ε3). In the sensitivity analyses, we constructed another continuous *APOE* score according to the odds ratios (ORs) in the meta-analysis of *APOE* genotype frequencies from AlzGene [26].

### 2.5 Statistical analyses

All analyses were done in R (version 4.2.1). To examine whether and how age and *APOE* ε4 interact with non-*APOE* PRS to influence cognitive decline, we modelled longitudinal standardized global and domain-specific cognitive trajectories using linear mixed-effect models (lme4 package, R). Models include PRS, *APOE* ε4, age, age^2^, their pairwise interactions, and three-way interactions between them (PRS × *APOE* ε4 × all polynomial terms of age), along with a set of covariates: sex, education years, parental history of AD, practice effects, and the first five principal components (to control for population stratification), as well as random intercept at both subject and family level to account for within-family (sibship) and within-subject correlations. We centered age at 65 and education level at the mean of all visits for ease of interpretation. The joint significance of the three-way interaction between PRS, *APOE* ε4, and polynomial age was tested using likelihood ratio tests between the full model defined above with and without the three-way interaction between PRS, *APOE* ε4, and age (PRS × *APOE* ε4 × age and PRS × *APOE* ε4 × age^2^). We next compared the model with the three-way interactions between PRS, *APOE* ε4, and polynomial age to the models that only considered the main effect of PRS and *APOE* ε4, only the *APOE* ε4 × age interaction, only the PRS × age interaction, only the PRS × *APOE* ε4 interaction, and the model that includes PRS × age and *APOE* ε4 × age but no PRS × *APOE* interaction. We used the Akaike information criterion-corrected (AICc; an AICc difference of 4 is considered to be meaningful, suggesting that a model with a lower AICc is a better fit [27]) statistics and likelihood ratio tests (LRT) for the goodness of fit between these models. Two-sided *P* values <0.05 were considered statistically significant. Upon the discovery of statistically significant three-way interactions, we further probed the nature of these interaction effects by conducting post-hoc simple slope analyses (reghelper and emmeans package, R). Specifically, we first calculated the simple slope estimates (conditional effect) of non-*APOE* PRS on longitudinal cognition for both *APOE* ε4 carriers and non-carriers from 55 to 80 years of age and tested them versus zero for investigating the threshold of significance. We next tested when and whether there is a statistically significant difference in the simple slope estimates of non-*APOE* PRS between *APOE* ε4 carriers and non-carriers at the same age. For all post-hoc analyses, we have used false discovery rate (FDR)-adjusted *P* value for significance to minimize the threats caused by multiple comparison issues (due to simultaneously testing the significance of simple slopes and the difference of simple slopes at various age points). Except PRS_Kunkle_sig_ in the main analysis, in the sensitivity analyses, we repeated the above analyses by replacing the PRS predictor constructed using different *P*-threshold or summary statistics (e.g., PRS_Kunkle_1e-5_, PRS_Kunkle_1e-3_, PRS_Kunkle_1e-1_, PRS_Kunkle_1_, PRS_deroja_sig_), and by replacing binary *APOE* ε4 carrier status with the continuous *APOE* score.

### 2.6 Replication analyses

We replicated our main WRAP analyses in the Health and Retirement Study (HRS), a national representative longitudinal biennial panel that has surveyed ∼42,000 Americans from 26,000 households since 1992. Genetic data were collected in a subsample of approximately ∼15,000 participants from 2006 to 2012[28]. Since 2000, HRS has collected consistent measures on cognition with a 27-point modified version of the Telephone Interview for Cognitive Status (TICSm) [29–32]. The TICSm assesses immediate recall (0-10 points) and delayed recall (0-10 points) tests for memory performance, serial 7s subtraction (0-5 points) tests of working memory, and backwards counting from 20 (0-2 points), which was designed to measure processing speed. Details about these tests have been described elsewhere [33]. A global cognition composite score was created by summing all the items within the tests mentioned above, with a maximum of 27 points. In the replication analysis, we used data from the 2000 wave of the HRS with follow-up until 2018. To ensure a fair comparison to the WRAP results, we focused on the preclinical stage of Alzheimer’s by only including HRS participants who were born between 1935 and 1959 (age 40 to 65 at year 2000), were non-Hispanic white, whose cognition was not assessed through proxy, and were not classified as “demented” by the Langa-Weir Classification of Cognitive Function [31].

Genotype data on over 15,000 HRS participants was obtained using the Illumina HumanOmni2.5 BeadChip[34]. Individuals with missing call rates > 2% or chromosomal anomalies were excluded. Also, SNPs that do not meet the quality control criteria, including intensity-only or duplicate SNPs, SNPs with MAF = 0, missing call rate ≥ 2%, > 2 discordant calls in 103 study duplicates, >1 Mendelian error, Hardy-Weinberg equilibrium p-value < 1e-4, sex difference in allelic frequency ≥ 0.2, and sex difference in heterozygosity > 0.3 were excluded. Genotype data were imputed to a worldwide 1000 Genomes Project reference panel using SHAPEIT2 [35] and IMPUTE2 [36] software. Genotype imputation was performed and documented by the Genetics Coordinating Center at the University of Washington. Only SNPs that were directly genotyped or imputed with info score > 0.8 were kept in the analysis. We replicated the WRAP main analyses by using PRS_Kunkle_sig_ as the main polygenic predictor and used the same procedure of constructing PRS as that described in the WRAP analyses.

WRAP findings on immediate learning, delayed recall, and PACC-3 were replicated on the HRS measures immediate recall, delayed recall, and global cognition, respectively. We could not replicate WRAP findings on executive function in HRS because neither serial 7s or counting backwards from 20 is quite like the tests included in the construction of the WRAP executive function score. All other statistical methods in the replication analyses were the same as those described in the WRAP analysis, except for the exclusion of parental history of AD as a covariate because measures on family history of AD were collected after 2010 [37]. We also included an indicator for cohort as a covariate because HRS enrolled a new cohort every six years and this measure was adjusted by the other AD-related HRS longitudinal cognition analyses [15]. To make the HRS replication analyses comparable to the WRAP findings, we additionally standardized all the cognitive outcomes with a mean of 0 and standard deviation of 1.

## 3. RESULTS

### 3.1 Descriptive statistics of the WRAP sample

We have presented the descriptive statistics of the WRAP sample when all measures for global and domain-specific cognition score first became available (visit 2) in Table 1. Briefly, a total of 1,190 individuals with available genetic, cognitive, and demographic data remained in the sample for up to six visits (about ten years) after the implementation of the inclusion criteria (see methods). The mean age at visit 2 was 58.6 years, the mean education was 15.8 years, about 30% of WRAP participants were male, about 73% had a parental history of AD, and about 39% of participants were *APOE* ε4 carriers.

**Table 1.** Descriptive statistics of WRAP sample at visit 2

| **Variables** | **N = 1,190** |
| --- | --- |
| Age at baseline<sup>1</sup> (Mean (SD)) | 58.6 (6.5) |
| Education (Mean (SD)) | 15.8 (2.3) |
| Family History of AD (n / N (%)) | 863 / 1,190 (73%) |
| Male (n / N (%)) | 361 / 1,190 (30%) |
| ***APOE* alleles (n / N (%))** |  |
| ε2/ε2 | 4 / 1,190 (0.3%) |
| ε2/ε3 | 101 / 1,190 (8%) |
| ε2/ε4 | 39 / 1,190 (3%) |
| ε3/ε3 | 626 / 1,190 (53%) |
| ε3/ε4 | 376 / 1,190 (32%) |
| ε4/ε4 | 44 / 1,190 (4%) |
| **Maximum Visit<sup>2</sup> (n / N (%))** |  |
| 1 | 44 / 1,190 (4%) |
| 2 | 84 / 1,190 (7%) |
| 3 | 149 / 1,190 (13%) |
| 4 | 297 / 1,190 (25%) |
| 5 | 444 / 1,190 (37%) |
| 6 | 172 / 1,190 (14%) |

<sup>1</sup> “Baseline age” refer to the age when cognition data first become available.
<sup>2</sup> “Maximum visits” refers to number of cognitive assessments of outcomes included in these analyses.

### 3.2 Comparison of main effect, two-way interaction, and three-way interaction models

We first compared the model fit between the three-way interaction model and a set of models that have been commonly investigated in previous LOAD genetic studies (see supplementary methods). This model set includes the main effect model, with only the main effects of *APOE*, PRS, and age [26,38]; adding only the interaction between *APOE* and age [7,8]; adding only the interaction between PRS and age [8,9]; adding only the interaction between PRS and *APOE* [11,13]; and the model that adds the age-related genetic effects of both PRS and *APOE* but no PRS × *APOE* interaction.

As shown in Table 2, for delayed recall, the best fitting model is the three-way interaction model between PRS_Kunkle_sig_, *APOE* ε4, and age, as this model has the lowest AICc statistics (AICc = 9979.36) compared to all other models. Results from LRTs also indicate the three-way interaction model significantly fits the data better than all other models. Similar findings were observed for PACC-3 but not for immediate learning and executive function.

**Table 2.**
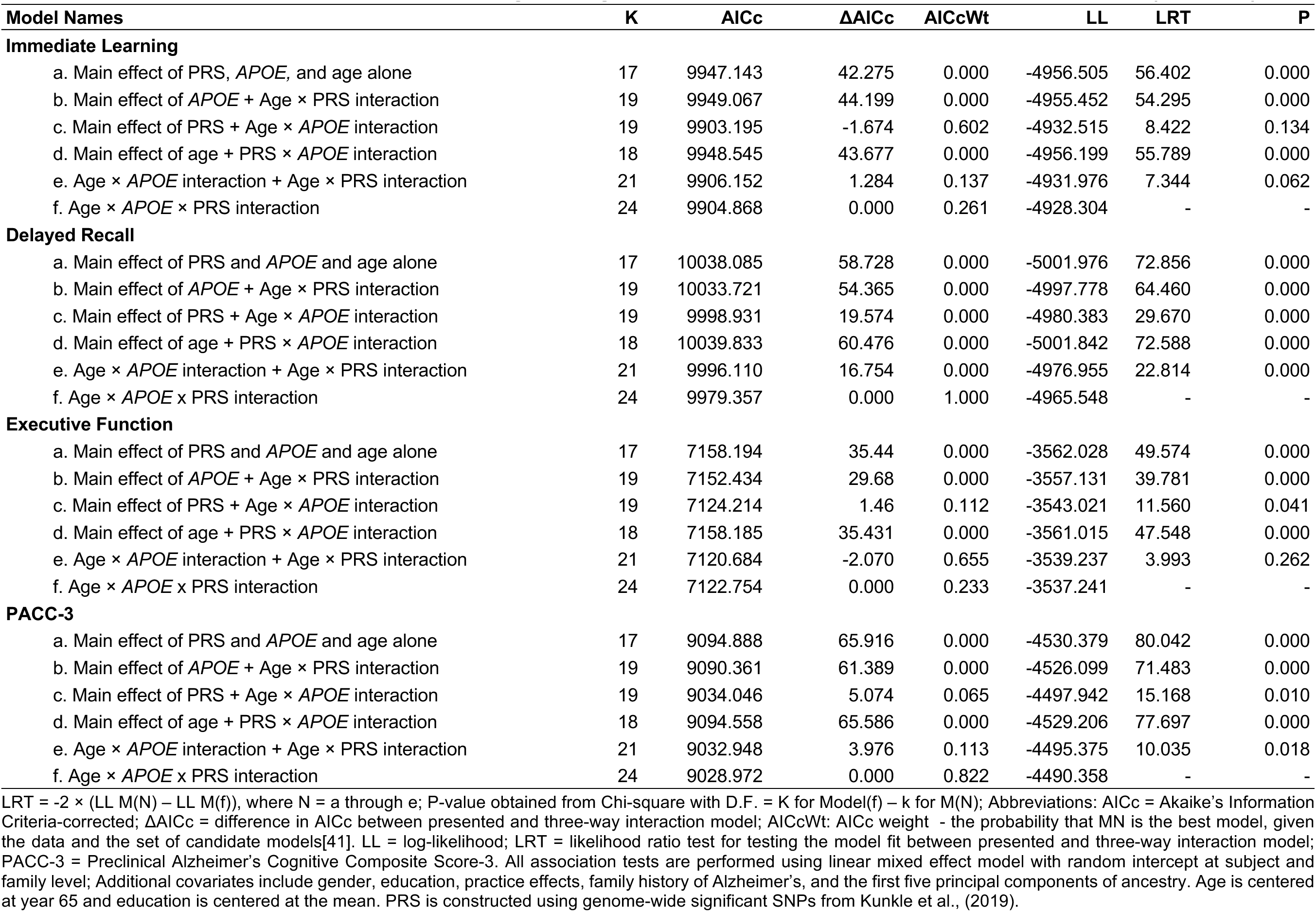
Model fit statistics, AICc, model weights, log-likelihood, and likelihood ratio tests in WRAP (N=1,190)

### 3.3 Interaction between PRS, *APOE* ε4, and age

Our primary interest is to understand whether *APOE* ε4 modifies the association between PRS and age on global and domain-specific cognitive performance. For delayed recall (Table 3), the PRS_Kunkle_sig_ × *APOE* ε4 × age interaction provided strong statistically significant evidence that *APOE* ε4 modifies PRS-related cognitive decline (PRS_Kunkle_sig_ × *APOE* ε4 × Age, *P* < 0.001; PRS_Kunkle_sig_ × *APOE* ε4 × Age^2^, *P* < 0.001). Similar findings were observed for immediate learning (PRS_Kunkle_sig_ × *APOE* ε4 × Age, *P* = 0.037; PRS_Kunkle_sig_ × *APOE* ε4 × Age^2^, *P* = 0.024) and PACC-3 (PRS_Kunkle_sig_ × *APOE* ε4 × Age, *P* = 0.02; PRS_Kunkle_sig_ × *APOE* ε4 × Age^2^, *P* = 0.027). However, neither linear nor quadratic terms of age significantly interacted with *APOE* ε4 and PRS_Kunkle_sig_ for executive function (PRS_Kunkle_sig_ × *APOE* ε4 × Age, *P* = 0.266; PRS_Kunkle_sig_ × *APOE* ε4 × Age^2^, *P* = 0.331). We next performed likelihood ratio tests to examine the joint significance of the three-way interaction with polynomial age terms for all cognitive outcomes. Statistically significant three-way interactions based on LRTs were observed for immediate learning (LRT = 6.563, DF = 2, *P* = 0.038), delayed recall (LRT = 22.331, DF = 2, *P* < 0.001), and PACC-3 (LRT = 7.264, DF = 2, *P* = 0.026). However, we failed to reject the null hypothesis that there is no PRS_Kunkle_sig_ × *APOE* ε4 × Age interaction on executive function (LRT = 1.518, DF = 2, *P* = 0.468).

**Table 3.** Associations of *APOE* ε4 carrier status and non-*APOE* PRS_Kunkle_sig_ with <u>global and domain specific longitudinal cognition score in WRAP (N=1,190)</u>

|  | Immediate Learning | Delayed Recall | Executive Function | PACC-3 |
| --- | --- | --- | --- | --- |
| (Intercept) | -0.7223 ***<br>(0.0683) | -0.6719 ***<br>(0.0697) | -0.7201 ***<br>(0.0659) | -0.8323 ***<br>(0.0665) |
| PRS | 0.0223<br>(0.0332) | 0.0423<br>(0.0339) | 0.0549 *<br>(0.0315) | 0.0323<br>(0.0322) |
| *APOE* ε4 | -0.1392 **<br>(0.0571) | -0.1230 **<br>(0.0583) | -0.1179 **<br>(0.0545) | -0.1765 ***<br>(0.0554) |
| Age | -0.0473 ***<br>(0.0045) | -0.0448 ***<br>(0.0045) | -0.0724 ***<br>(0.0041) | -0.0603 ***<br>(0.0043) |
| Age<sup>2</sup> | -0.0004 **<br>(0.0002) | -0.0006 ***<br>(0.0002) | -0.0009 ***<br>(0.0002) | -0.0008 ***<br>(0.0002) |
| PRS × Age | 0.0012<br>(0.0027) | 0.0035<br>(0.0027) | -0.0035<br>(0.0022) | -0.0005<br>(0.0024) |
| PRS × *APOE* ε4 | -0.0269<br>(0.0556) | -0.0115<br>(0.0567) | -0.0758<br>(0.0522) | -0.0724<br>(0.0539) |
| *APOE* ε4 × Age | -0.0291 ***<br>(0.0047) | -0.0267 ***<br>(0.0047) | -0.0215 ***<br>(0.0037) | -0.0307 ***<br>(0.0041) |
| PRS × Age<sup>2</sup> | 0.0001<br>(0.0002) | -0.0001<br>(0.0002) | -0.0002<br>(0.0002) | -0.0001<br>(0.0002) |
| *APOE* ε4 × Age<sup>2</sup> | -0.0020 ***<br>(0.0004) | -0.0020 ***<br>(0.0004) | -0.0011 ***<br>(0.0003) | -0.0017 ***<br>(0.0003) |
| PRS × *APOE* ε4 × Age | -0.0100 **<br>(0.0048) | -0.0200 ***<br>(0.0049) | -0.0042<br>(0.0038) | -0.0098 **<br>(0.0042) |
| PRS × *APOE* ε4 × Age<sup>2</sup> | -0.0009 **<br>(0.0004) | -0.0015 ***<br>(0.0004) | -0.0003<br>(0.0003) | -0.0008 **<br>(0.0003) |

### 3.4 Post-hoc simple slope analyses

We performed post-hoc simple slope analyses to probe the nature of the significant PRS_Kunkle_sig_ × *APOE* ε4 × polynomial age interactions that we have identified in the above analysis (Figure 1 and Figure 2, top panel within each outcome, and Supplementary Table 1). First, we tested each of the simple slopes of PRS_Kunkle_sig_ on all cognitive outcomes among people with and without *APOE* ε4, from age 55 to 80, for significance. For delayed recall, after FDR correction, statistically significant simple slope estimates of PRS_Kunkle_sig_ emerged after people reached age 72 (*P_FDR_* = 0.017) among *APOE* ε4 carriers, and the simple slope absolute effect size accelerated in growth thereafter and remained statistically significant. However, we did not observe any statistically significant simple slope estimates of PRS_Kunkle_sig_ before *APOE* ε4 carriers reached age 72. Among *APOE* ε4 non-carriers, we did not observe any statistically significant simple slope estimates of PRS_Kunkle_sig_ at any ages we tested. Similar findings were observed on immediate learning and PACC-3. Among *APOE* ε4 carriers, we observed that statistically significant simple slope estimates of PRS_Kunkle_sig_ emerged after people reached age 70 for PACC-3 (*P_FDR_*=0.048) and borderline statistically significant simple slope estimates of PRS_Kunkle_sig_ emerged after people reached age 75 (*P_FDR_* = 0.083) for immediate learning. Among *APOE* ε4 non-carriers, we did not observe any statistically significant simple slope estimates of PRS_Kunkle_sig_ on either PACC-3 or immediate learning at any ages we tested. For executive function, we did not observe any statistically significant simple slope estimates of PRS_Kunkle_sig_ after FDR correction, regardless of age or *APOE* ε4 carrier status.

**Figure 1.**
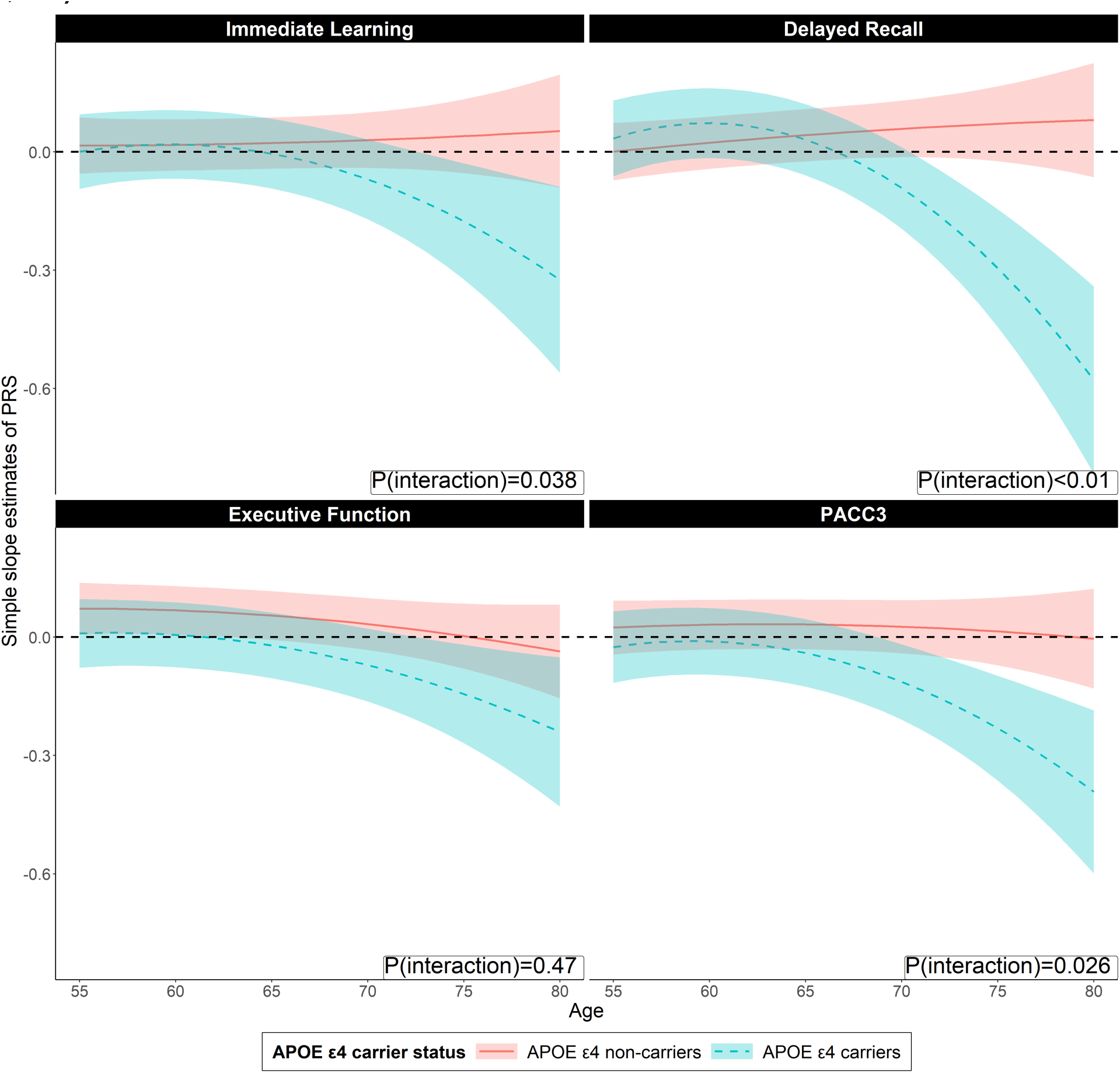
Simple slope estimates of PRS_Kunkle_sig_ on domain specific– and global cognitive score for individuals with and without *APOE* ε4 and at different age (N = 1,190). Figure 1 shows the simple slope estimates of the PRS for individuals with and without *APOE* ε4 from age 55 to 80 on global and domain specific cognition score. The red line represents the longitudinal trajectory of simple slope estimates of PRS_Kunkle_sig_ among *APOE* ε4 non-carriers while the blue line represents simple slope estimates of PRS_Kunkle_sig_ among individuals with *APOE* ε4. Bands represent 95% Confidence intervals. The simple slope estimates are calculated using the package “reghelper” in R and were based on the results which were obtained using the linear mixed-effect model and adjusted for within-individual/family correlation. In addition to PRS, age (quadratic), *APOE* ε4, and their interactions, additional covariates include gender, education years, practice effect, parental history of AD, and the first five principal components of ancestry. Age is centered at year 65 and education is centered at the mean. PACC-3 = Preclinical Alzheimer’s Cognitive Composite Score-3.

**Figure 2.**
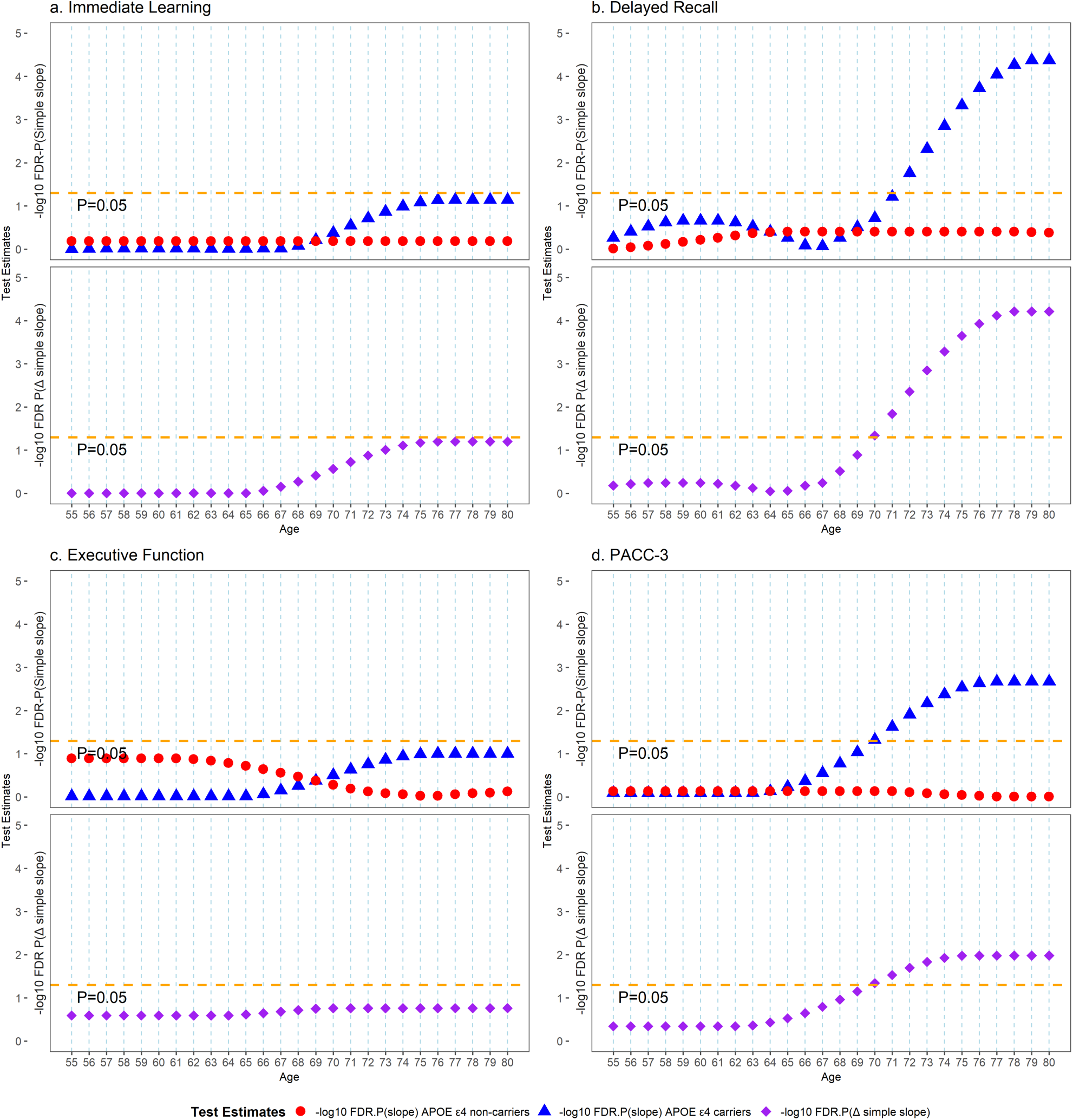
FDR-corrected *P*-value of simple slope estimates of PRS_Kunkle_sig_ and difference in simple slope estimates on domain specific– and global cognitive score for individuals with and without *APOE* ε4 and at different ages (N = 1,190). Figure 2 shows the longitudinal trajectory of the change in the significance of the simple slope estimates of PRS for people with and without *APOE* ε4 but at different ages. Within each outcome, the top panel represents the longitudinal trajectory of the change in significance (after FDR correction) of the simple slope estimates for people with and without *APOE* ε4 but at different ages. For example, the red point represents the FDR-corrected p-value of the simple slope estimates of PRS among *APOE* ε4 non-carriers at various ages. In contrast, the blue point represents the FDR-corrected p-value of the simple slope estimates of PRS among *APOE* ε4 carriers at different ages. The bottom panel represents the longitudinal trajectory of the change in significance (after FDR correction) of the difference in simple slopes for people with and without *APOE* ε4 but at the same age as people become older. For example, the purple point represents the FDR-corrected p-value of the difference in simple slope estimates of PRS between *APOE* ε4 carriers and non-carriers at various ages. The reference line in the top and bottom panel represents P-value of simple slope of PRS =0.05, P-value of simple slope of PRS between ε4 carrier/ ε4 non-carrier=0.05, respectively. PACC-3 = Preclinical Alzheimer’s Cognitive Composite Score-3.

We subsequently tested whether there is a statistically significant difference in the simple slope estimates of PRS_Kunkle_sig_ on cognition between individuals of the same age with and without *APOE* ε4 at a range of ages (Figure 2 bottom panel within each outcome and Supplementary Table 2). The model parameters predicted that significant PRS_Kunkle_sig_-related differences between people with and without *APOE* ε4 started showing once people reached age 70 for delayed recall (*P_FDR_*=0.046) and PACC-3 (*P_FDR_*=0.045). Borderline significant PRS_Kunkle_sig_-related differences between people with and without *APOE* ε4 were observed for immediate learning once people reached age 74 (*P_FDR_*=0.078), but no statistically significant differences in the simple slope estimates were observed for executive function at any age investigated.

### 3.5 Sensitivity analyses

We next tested PRS_deroja_sig_ and IGAP’s GWAS-informed PRSs with different *P* value thresholds in the three-way interaction model to check the consistency of the results (Figure 3, Supplementary Tables 1–5). In terms of model comparison based on AICc and LRT statistics, the PRS_deroja_sig_ × *APOE* ε4 × age interaction model outweighed all other models (see methods) on immediate learning, delayed recall, and PACC-3. Borderline statistically significant evidence showed that the three-way interaction model with PRS_Kunkle_1e-5_/PRS_Kunkle_1e-3_ was better than all other models on delayed recall and PACC-3. When we used PRS_Kunkle_1e-1_/PRS_Kunkle_1_ as the predictor, the three-way interaction model was no longer the best model for all cognitive outcomes. For immediate learning, statistically significant PRS × *APOE* ε4 × age interactions were only observed on PRS_deroja_sig_. For delayed recall and PACC-3, statistically significant PRS × *APOE* ε4 × age interactions were observed when we used PRS_deroja_sig_ and constructed PRS using IGAP’s GWAS with a conservative *P* value threshold (e.g., *P* < 1e^-5^), but we did not observe any statistically significant PRS × *APOE* ε4 × age interaction on these two cognitive outcomes when we used a PRS with a relatively liberal *P* value threshold (e.g., *P* < 0.1 and *P* < 1). Surprisingly, for executive function, we only observed a statistically significant PRS × *APOE* ε4 × age interaction when we used a liberal *P* value threshold (*P* < 1e^-3^, *P* < 0.1, and *P* < 1). In terms of simple slope analysis (Supplementary Figures 1–4, Supplementary Tables 1 and 2), the age-related trajectory of the PRS effect is the most similar to that reported in the main results when we used PRS_deroja_sig_ as the predictor. When we used PRS_Kunkle_1e-5_ or PRS_Kunkle_1e-3_ as the predictor, the results for the age-related trajectory of the PRS effects were also similar to the main results, but the age threshold of the significant simple slope estimates among *APOE* ε4 carriers and the age threshold of the significant PRS-related difference became older on immediate learning, delayed recall, and PACC-3. For PRS_Kunkle_1e-1_ and PRS_Kunkle_1_, no statistically significant simple slope estimates or difference were observed on immediate learning, delayed recall, and PACC-3 at all ages. Results for replacing the binary *APOE* ε4 status with a continuous *APOE* score in the three-way interaction model were similar to the main study results (Supplementary Figure 5, Supplementary Tables 3–5 and 6–7).

**Figure 3.**
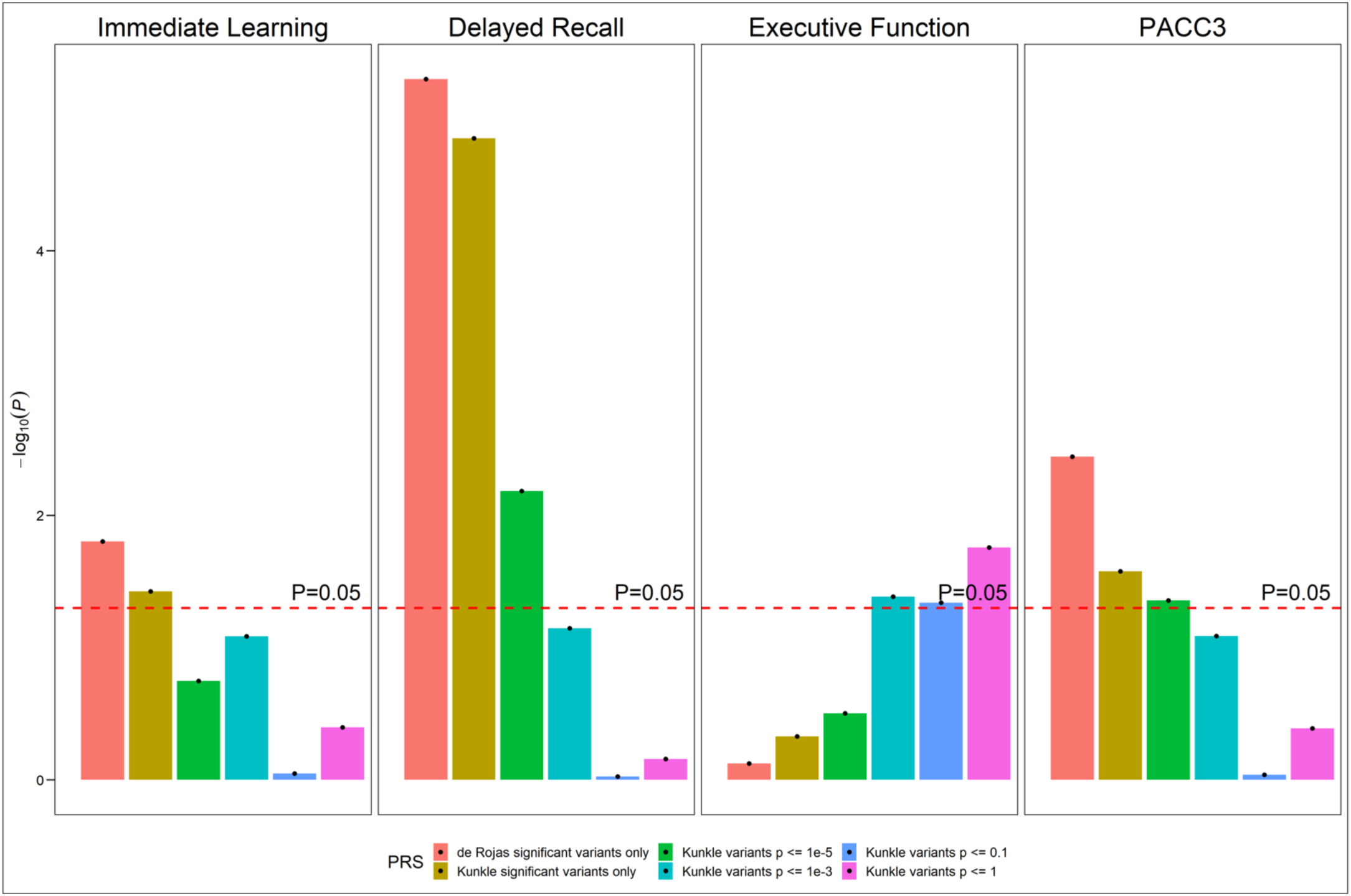
Likelihood Ratio Test (LRT) of the interactions between different PRSs with different *P*-thresholds, *APOE*-ε4 status, and Age. Figure 3 presents the –log_10_(*P*) from the likelihood ratio tests for the three-way interaction terms for all outcomes and different PRSs in WRAP. The likelihood ratio test statistic is calculated as the ratio between the log-likelihood of the nested model (model without three-way interaction terms) to the full model (model with polynomial age*PRS\**APOE* terms). All association tests are performed using linear mixed effect model with random intercept at subject and family level; Additional covariates include gender, education, practice effects, family history of Alzheimer’s, and the first five principal components of ancestry. Age is centered at year 65 and education is centered at the mean. PACC-3 = Preclinical Alzheimer’s Cognitive Composite Score-3.

### 3.6 Replication analyses

We have presented the descriptive statistics of the HRS analytical sample in Supplementary Table 8. Briefly, a total of 6,785 HRS participants remained in the study after the implementation of the inclusion criteria. The mean age at enrollment was 56.5, the mean education was 13.5 years, about 57% of HRS participants were female, and about 26% of participants were *APOE* ε4 carriers. Key findings in WRAP were successfully replicated in the HRS analytical sample. In terms of model comparison based on AICc and LRT statistics, the PRS_Kunkle_sig_ × *APOE* ε4 × age interaction model outperformed all other models on immediate recall, delayed recall, and global cognition (Supplementary Table 3. Statistically significant interactions between polynomial age, *APOE* ε4, and PRS_Kunkle_sig_ were observed on all cognitive outcomes (Supplementary Table 4). The results for the simple slope analyses were also similar to the WRAP findings. Specifically, after FDR correction and among *APOE* ε4 carriers, statistically significant simple slope estimates of PRS_Kunkle_sig_ were observed to be negatively associated with delayed recall and global cognition after age 66 and with immediate recall after age 69 (Supplementary Figure 6, Supplementary Table 9). Among *APOE* ε4 non-carriers, we did not observe any statistically significant simple slope estimates of PRS_Kunkle_sig_ on any cognitive outcomes at any ages in HRS. In terms of the difference in the simple slope estimates, the model parameters predicted that significant PRS_Kunkle_sig_-related differences between people with and without *APOE* ε4 started showing after people reached age 67 on global cognition, after 69 on delayed recall, and after 73 on immediate recall (Supplementary Table 10). We performed a parallel analysis in WRAP by excluding the parental history of AD as a covariate, and the results remained unchanged.

## 4. DISCUSSION

In this study, we investigated whether *APOE* ε4 carrier status can modify the age-related effect of non-*APOE* PRS on longitudinal global and domain-specific cognition in a group of cognitively unimpaired, late-middle-aged Wisconsin adults enriched for parental history of AD. We also replicated our focal study findings in a sample of non-demented individuals with a similar age range from a nationwide population-based longitudinal study. We evaluated the robustness of the association between non-*APOE* PRS x *APOE* x age by referring to summary statistics from different recent GWAS meta-analyses to construct PRS, varying the functional form of *APOE* and including different numbers of SNPs in the polygenic predictor. We found a statistically significant association between non-*APOE* PRS x *APOE* x age and longitudinal cognition. The adverse genetic effect caused by non-*APOE* PRS is more substantial among *APOE* ε4 carriers after people reach age 70. These associations are stronger when we construct non-*APOE* PRS using a conservative *P*-value selection (e.g., *P* < 5e^-8^) threshold but dissipate as we increase the threshold (e.g., *P* < 1). To our knowledge, this is the first study that thoroughly examined the non-*APOE* PRS x *APOE* x age interaction on longitudinal cognition and with a particular focus on the preclinical stage of Alzheimer’s.

Previous studies have reported mixed results regarding the modifying effect of *APOE* ε4 on the association between PRS and AD-related phenotypes. Our study findings are consistent with the population-based Rotterdam Study that the effect of PRS on AD and dementia is more substantial among *APOE* ε4 carriers than non-carriers[11]. Other studies also observed the significant effect of PRS in *APOE* ε4 carriers when they focused on AD-related biomarkers or progression to AD[10,12]. Our findings are also consistent with Fulton-Howard et al. [25], who tested the interaction between PRS x *APOE* x age and found that known AD risk variants are particularly detrimental in *APOE* ε4 carriers. Our findings are inconsistent with Najar et al. [13], who assessed the modifying effect of *APOE* ε4 in the oldest old and found an association between PRS and dementia among *APOE* ε4 non-carriers. As discussed in their paper, one reason for the discrepancy in results could be the sample age, with a mean age at baseline of 80 years. This inclusion of older participants could lead to healthier *APOE* ε4 carriers who may carry additional protective genetic variants that prevent them from developing dementia. In the HRS, we repeated the primary analysis by lifting birth year restrictions (1910-1959). Even though we still observed a more significant effect of PRS among *APOE* ε4 carriers, the PRS effect and the difference in the PRS effect between *APOE* ε4 carriers and non-carriers attenuated substantially compared to when we ran the analyses in the younger sample (results not shown).

Consistent with Najar et al., no statistically significant interactions were found between *APOE* ε4 carriership and PRS when the PRS was constructed using a more liberal *P*-value selection threshold for SNPs (e.g., *P* < 1e^-3^), even after considering the age-related genetic effect. In HRS, Ware et al. also failed to observe statistically significant interactions between *APOE* and PRS on dementia when constructing the PRS using a *P*-threshold of 1e^-2^[15]. There is no consensus on which *P-*threshold is optimal for polygenic prediction models in AD research. Most current research has found statistically significant modifying effects of *APOE* on a PRS constructed using a conservative *P-*threshold. Although Escott Price et al. reported that the PRS including SNPs with a *P*-threshold of 0.5 had the highest prediction of AD[39], a liberal approach for *P*-threshold selection could also introduce non-informative SNPs and limit the discriminative ability of the genetic predictor[40].

In our previous WRAP analyses, without considering the PRS x *APOE* x age interaction, we found that the effect of the non-*APOE* PRS starts emerging on preclinical cognition decline after people reach age 75. When we combined PRS, *APOE,* and age, we discovered that non-*APOE* PRS’s effect started appearing on preclinical cognition around age 70 for delayed recall and PACC-3 among *APOE* ε4 carriers. Among *APOE* ε4 non-carriers, we did not observe any statistically significant effects of PRS on all cognitive outcomes at all ages we have investigated. Replication results in the population-based HRS also supported these findings. These findings are consistent with previous studies that non-*APOE* PRS has greater role in modifying the age-at-onset among *APOE* ε4 carriers than non-carriers[11,25]. The associations between PRS x *APOE* x age are more pronounced on immediate learning, delayed recall, and PACC-3 than on executive function. One possible reason is that STROOP tests in WRAP were dropped in 2019 so the executive function composite score was unavailable for more recent visits. The exclusion of the longitudinal follow-up of this measure in later ages could attenuate the association between PRS x *APOE* x age and executive function. Even though we observed a statistically significant PRS × *APOE* ε4 × age interaction from LRT test when we used a liberal *P* value threshold (*P* < 1e^-3^, *P* < 0.1, and *P* < 1) for executive function. we didn’t observe that the model parameters predicted that significant PRS related differences would emerge in the age range we have investigated when the PRS were constructed using these liberal *P* value thresholds (results not shown, available upon request).

The more significant effect of PRS on cognition among *APOE* ε4 carriers, as reflected by the increasing magnitude of the PRS effect size and the younger age threshold when a statistically significant PRS effect appears in the simple slope analyses, suggests the combined effect of currently known non-*APOE* genetic variants is more detrimental among *APOE* ε4 carriers. Similar to the literature, this finding implies the interaction might be additive, as individuals who are *APOE* ε4 carriers with high PRS are the most vulnerable to the harmful effects caused by genetic burden[12]. The findings also mean an increased risk of *APOE* ε4 can be mitigated by having a low PRS. Even though PRS is not currently applied in clinical settings for risk profiling, our study shows that the genetic risk of Alzheimer’s can be more comprehensively estimated by integrating both *APOE* and PRS. Our analysis also provides insights into precision medicine by helping identify groups of people with an elevated risk of AD at a younger age in future clinical trials, as the selection of those from the highest-risk groups in the clinical trials can substantially reduce the associated costs and shorten the duration of the trial. Our study’s findings can serve as the foundation for future epidemiological studies, which focus on identifying protective factors that can mitigate the genetic risk of Alzheimer’s, especially gene-environment interaction analyses.

Our study has some limitations. First, we only focus on the preclinical cognitive function as the outcome but have not extended the study to AD-related biomarkers because of the concern for the balance between model complexity and sample size (biomarker samples usually have small sample sizes, N < 200). The absence of information from the study of those endophenotypes limits our ability to interpret the biological nature behind the observed interaction effects. Second, WRAP reflects the demographics of Wisconsin adults, and the findings’ generalizability are limited. Even though we replicated the analyses in the population-based HRS by focusing on the individuals who have not reached the “demented” stage, the dementia status classified in HRS is purely based on the cognitive score and the percentage of “misdiagnoses” might be high. Third, it is essential to note that our study findings might not be directly generalizable to the population other than those of European descent. As larger non-European ancestry GWAS are published, providing summary statistics for PRS calculation, additional analyses will be needed to include people from diverse ancestral backgrounds to better understand the general population’s non-*APOE* PRS x *APOE* x age interaction.

In summary, we found a statistically significant association between non-*APOE* PRS x *APOE* x age and longitudinal cognition in a convenience sample enriched for individuals with a parental history of AD. We replicated our study findings in a large population-based sample. The adverse genetic effect caused by currently known common genetic variants is more detrimental among *APOE* ε4 carriers once they reach age 70. Our findings contribute to the efficacy of future clinical trials and provide insights for future epidemiological studies to identify protective factors that can mitigate the adverse effects caused by genetic risk factors, such as gene-environment interaction analyses in Alzheimer’s.

## Data Availability

All data produced in the present study are available upon reasonable request to the authors.

## ACKNOWLEDGEMENTS

The authors especially thank the WRAP participants and staff for their contributions to the studies. Without their efforts, this research would not be possible. This study was supported by the National Institutes of Health (NIH) grants R01AG27161 (Wisconsin Registry for Alzheimer Prevention: Biomarkers of Preclinical AD), R01AG054047 (Genomic and Metabolomic Data Integration in a Longitudinal Cohort at Risk for Alzheimer’s Disease), and R21AG067092 (Identifying Metabolomic Risk Factors in Plasma and Cerebrospinal Fluid for Alzheimer’s Disease), the Helen Bader Foundation, Northwestern Mutual Foundation, Extendicare Foundation, State of Wisconsin, the Clinical and Translational Science Award (CTSA) program through the NIH National Center for Advancing Translational Sciences (NCATS) grant [UL1TR000427]. Computational resources were supported by core grants to the Center for Demography and Ecology [P2CHD047873] and the Center for Demography of Health and Aging [P30AG017266]. Author YX was supported by a Center for Demography of Health and Aging pilot award [P30AG017266]. The HRS (Health and Retirement Study) is sponsored by the National Institute on Aging (grant number NIA U01AG009740) and is conducted by the University of Michigan.

## CONFLICTS OF INTEREST

SCJ has served as a consultant to Roche Diagnostics, Merck and Prothena and has received research funding from Cerveau Technologies.

## DATA AVAILABILITY

The WRAP data supporting the findings of this study are available through a resource request: https://wrap.wisc.edu/data-requests-2/. The data are not publicly available due to privacy and ethical restrictions. The HRS genetic and epigenetic data can be accessed via the National Institute on Aging Genetics of Alzheimer’s Disease Data Storage Site (NIAGADS, accession number: NG00119.v1). HRS cognition and survey data are publicly available from the HRS website at https://hrs.isr.umich.edu/data-products (accessed on 01 Janauary 2023).

## Notes

### Author Declarations

IRB of University of Wisconsin-Madison gave ethical approval for this work.

