## supplementary_methods_figures for "Apolipoprotein E moderates the association between Non-*APOE* Polygenic Risk Score for Alzheimer’s Disease and Aging on Preclinical Cognitive Function"

### Supplementary methods, tables, and figures

**Supplementary tables 1-12 see excel file**

**Supplementary methods**

**Supplementary figure 1.** Simple slope estimates of PRSs constructed using IGAP GWAS summary statistics with different thresholding parameters on immediate learning cognitive score for individuals with and without *APOE* ε4 and at different age (N = 1,190).

**Supplementary figure 2.** Simple slope estimates of PRSs constructed using IGAP GWAS summary statistics with different thresholding parameters on delayed recall cognitive score for individuals with and without *APOE* ε4 and at different age (N = 1,190).

**Supplementary figure 3.** Simple slope estimates of PRSs constructed using IGAP GWAS summary statistics with different thresholding parameters on executive function cognitive score for individuals with and without *APOE* ε4 and at different age (N = 1,190).

**Supplementary figure 4.** Simple slope estimates of PRSs constructed using IGAP GWAS summary statistics with different thresholding parameters on PACC-3 cognitive score for individuals with and without *APOE* ε4 and at different age (N = 1,190).

**Supplementary figure 5.** Simple slope estimates of $PRS_{Kunkle\_sig}$ on domain specific- and global cognitive score for individuals with different *APOE* genotype and at different age (N = 1,190).

**Supplementary figure 6.** Simple slope estimates of $PRS_{Kunkle\_sig}$ on domain specific- and global cognitive score for individuals with and without *APOE* ε4 and at different age in Health and Retirement Study (N = 6,785).

#### Supplementary methods

##### 1. Simple slopes calculation

Suppose we have a three-way interaction model

$Y= \beta_0 + \beta_1 PRS + \beta_2 Age + \beta_3 Age^2 + \beta_4 APOE + \beta_5 Age*PRS + \beta_6 Age^2*PRS + \beta_7 Age*APOE + \beta_8 Age^2*APOE + \beta_9 PRS*APOE + \beta_{10} PRS*APOE*Age + \beta_{11} PRS*APOE*Age^2 + \mathbf{BX} + i + f + u$

Where ***BX*** represents covariates (gender, education, parental history of AD, practice effects, and first five principal components), i index the within-individual random intercept, j is the within-family random intercept, and u is the error term.

To calculate the simple slope of the ***PRS*** for individuals with different *APOE* ε4 carrier status at different age, we can reorganize the above model as

$Y= \beta_0 + PRS(\beta_1 + \beta_5 Age + \beta_6 Age^2 + \beta_9 APOE + \beta_{10} APOE*Age + \beta_{11} APOE*Age^2) + \beta_2 Age + \beta_3 Age^2 + \beta_4 APOE + \beta_7 Age*APOE + \beta_8 Age^2*APOE + \mathbf{BX} + i + f + u$

Then it's easy to calculate the simple slope of PRS by using different combination of values for age and *APOE* ε4 in this three-way interaction model.

##### Example

If we want to calculate the simple slope of $PRS_{Kunkle\_sig}$ on PACC-3 for individuals with and without *APOE* ε4 and at age 65, 70, and 75.

We first get the regression outputs as the following (the same as the model presented in table 3, age is centered at year 65)

| | PACC-3 | | |
|---|---|---|---|
| *Predictors* | *Estimates* | *std. Error* | *p* |
| (β0) (Intercept) | -0.832294 | 0.066533 | **<0.001** |
| (β1) PRS | 0.032291 | 0.032169 | 0.316 |
| (β2) Age | -0.060302 | 0.004255 | **<0.001** |
| (β3) $Age^2$ | -0.000786 | 0.000190 | **<0.001** |
| (β4) APOE ε4 | -0.176490 | 0.055365 | **0.001** |
| (β5) PRS * Age | -0.000497 | 0.002370 | 0.834 |
| (β6) PRS * $Age^2$ | -0.000128 | 0.000179 | 0.475 |

| | | | |
|---|---|---|---|
| (β7) APOE ε4 * Age | -0.030727 | 0.004115 | **<0.001** |
| (β8) APOE ε4 * $Age^2$ | -0.001662 | 0.000316 | **<0.001** |
| (β9) PRS * APOE ε4 | -0.072353 | 0.053890 | 0.179 |
| (β10) PRS * Age * APOE ε4 | -0.009792 | 0.004199 | **0.020** |
| (β11) PRS * $Age^2$ * APOE ε4 | -0.000751 | 0.000340 | **0.027** |
| Female | 0.678827 | 0.053277 | **<0.001** |
| Years of education | 0.128226 | 0.011200 | **<0.001** |
| Practice effects | 0.107708 | 0.011031 | **<0.001** |
| Family history of AD | -0.040554 | 0.058385 | 0.487 |
| PC1 | -1.764619 | 0.847440 | **0.037** |
| PC2 | 0.477825 | 0.827262 | 0.564 |
| PC3 | 0.166100 | 0.840462 | 0.843 |
| PC4 | 0.083924 | 0.830247 | 0.919 |
| PC5 | 1.919996 | 0.827682 | **0.020** |

The simple slopes of $PRS_{Kunkle\_sig}$ on PACC-3 for individuals who are *APOE* ε4 non-carriers and at age 65, 70, 75 (0, 5, 10 for age-65-centered age) are

Age 65:

$$0.032291 + \beta_5 * 0 + \beta_6 * 0 + \beta_9 * 0 + \beta_{10} * 0 + \beta_{11} * 0 = 0.032291$$

Age 70:

$$0.032291 + (-0.000497) * 5 + (-0.000128) * 25 + \beta_9 * 0 + \beta_{10} * 0 + \beta_{11} * 0 = 0.026606$$

Age *75:*

$$0.032291 + (-0.000497) * 10 + (-0.000128) * 100 + \beta_9 * 0 + \beta_{10} * 0 + \beta_{11} * 0 = 0.014521$$

The simple slopes of $PRS_{Kunkle\_sig}$ on PACC-3 for individuals who are *APOE* ε4 carriers and at age 65, 70, 75 (0, 5, 10 for age-65-centered age) are

Age 65:

$$0.032291 + \beta_5 * 0 + \beta_6 * 0 + (-0.072353) * 1 + \beta_{10} * 0 + \beta_{11} * 0 = -0.040062$$

*Age 70:*

0.032291 + *(-0.000497)*5* + *(-0.000128)*25* + *(-0.072353)*1* + (-0.009792)**1*5* + (-0.000751)**1*25* = *-0.113482*

*Age 75:*

0.032291 + *(-0.000497)*10* + *(-0.000128)*100* + *(-0.072353)*1* + (-0.009792)**1*10* + (-0.000751)**1*100* = *-0.230852*

Visualize simple slopes in the predicted value plot

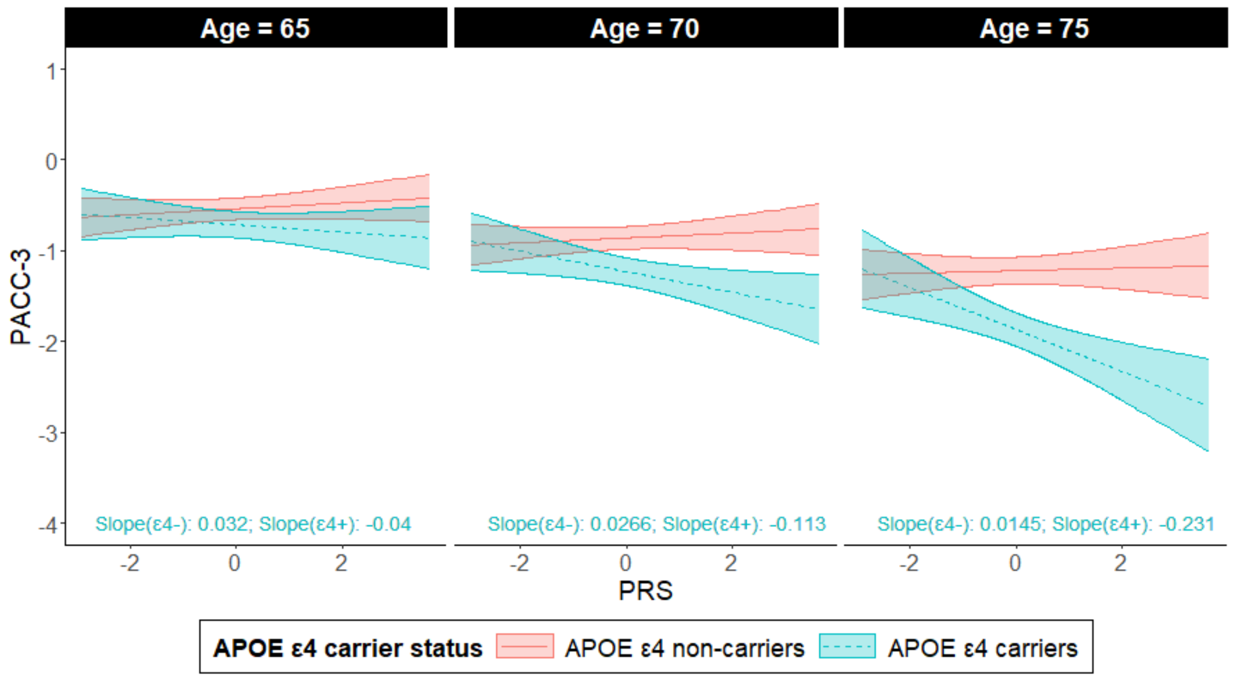

**2. Model set for model comparison**

*a. Main effect model*

$Y = \beta_0 + \beta_1 PRS + \beta_2 Age + \beta_3 Age^2 + \beta_4 APOE + \mathbf{BX} + i + f + u$ *(17 terms)*

*b. Main effect of APOE + Age × PRS interaction*

$Y = \beta_0 + \beta_1 PRS + \beta_2 Age + \beta_3 Age^2 + \beta_4 APOE + \beta_5 PRS^* Age + \beta_6 PRS^* Age^2 + \mathbf{BX} + i + f + u$ *(19 terms)*

*c. Main effect of PRS + Age × APOE interaction*

$Y = \beta_0 + \beta_1 PRS + \beta_2 Age + \beta_3 Age^2 + \beta_4 APOE + \beta_5 APOE^* Age + \beta_6 APOE^* Age^2 + \mathbf{BX} + i + f + u$ *(19 terms)*

*d. Main effect of age + PRS × APOE interaction*

$Y = \beta_0 + \beta_1 PRS + \beta_2 Age + \beta_3 Age^2 + \beta_4 APOE + \beta_5 PRS^* APOE + \mathbf{BX} + i + f + u$ *(18 terms)*

*e. Age × APOE interaction + Age × PRS interaction*

$Y = \beta_0 + \beta_1 PRS + \beta_2 Age + \beta_3 Age^2 + \beta_4 APOE + \beta_5 APOE^* Age + \beta_6 APOE^* Age^2 + \beta_7 PRS^* Age + \beta_8 PRS^* Age^2 + \mathbf{BX} + i + f + u$ *(21 terms)*

*f. Full model*

$Y = \beta_0 + \beta_1 PRS + \beta_2 Age + \beta_3 Age^2 + \beta_4 APOE + \beta_5 Age^*PRS + \beta_6 Age^{2*}PRS + \beta_7 Age^*APOE + \beta_8 Age^{2*}APOE + \beta_9 PRS^*APOE + \beta_{10} PRS^*APOE^*Age + \beta_{11} PRS^*APOE^*Age^2 + \mathbf{BX} + i + f + u$ *(24 terms)*

Where ***BX*** represents covariates (gender, education, family history of AD, practice effects, and first five principal components), i index the within-individual random intercept, j is the within-family random intercept, and u is the error term.

##### *3. Likelihood ratio tests*

We test the joint significance of the interactions between PRS, *APOE* ε4, and age using likelihood ratio tests. Specifically, we tested the coefficients for all the three-way interaction terms are simultaneously zero (degree of freedom = difference between number of parameters in the full model vs nested model).

Full model:

$Y = \beta_0 + \beta_1 PRS + \beta_2 Age + \beta_3 Age^2 + \beta_4 APOE + \beta_5 Age*PRS + \beta_6 Age^2*PRS + \beta_7 Age*APOE + \beta_8 Age^2*APOE + \beta_9 PRS*APOE + \beta_{10} PRS*APOE*Age + \beta_{11} PRS*APOE*Age^2 + \mathbf{BX} + i + f + u$ *(24 terms)*

Nested model:

$Y = \beta_0 + \beta_1 PRS + \beta_2 Age + \beta_3 Age^2 + \beta_4 APOE + \beta_5 PRS*APOE + \beta_6 APOE*Age + \beta_7 APOE*Age^2 + \beta_8 PRS*Age + \beta_9 PRS*Age^2 + \mathbf{BX} + i + f + u$ *(22 terms)*

Where ***BX*** represents covariates (gender, education, family history of AD, practice effects, and first five principal components), i index the within-individual random intercept, j is the within-family random intercept, and u is the error term.

**Supplementary figures 1. Simple slope estimates of PRSs constructed using IGAP GWAS summary statistics with different thresholding parameters on immediate learning cognitive score for individuals with and without *APOE* ε4 and at different age (N = 1,190).**

#### Immediate Learning

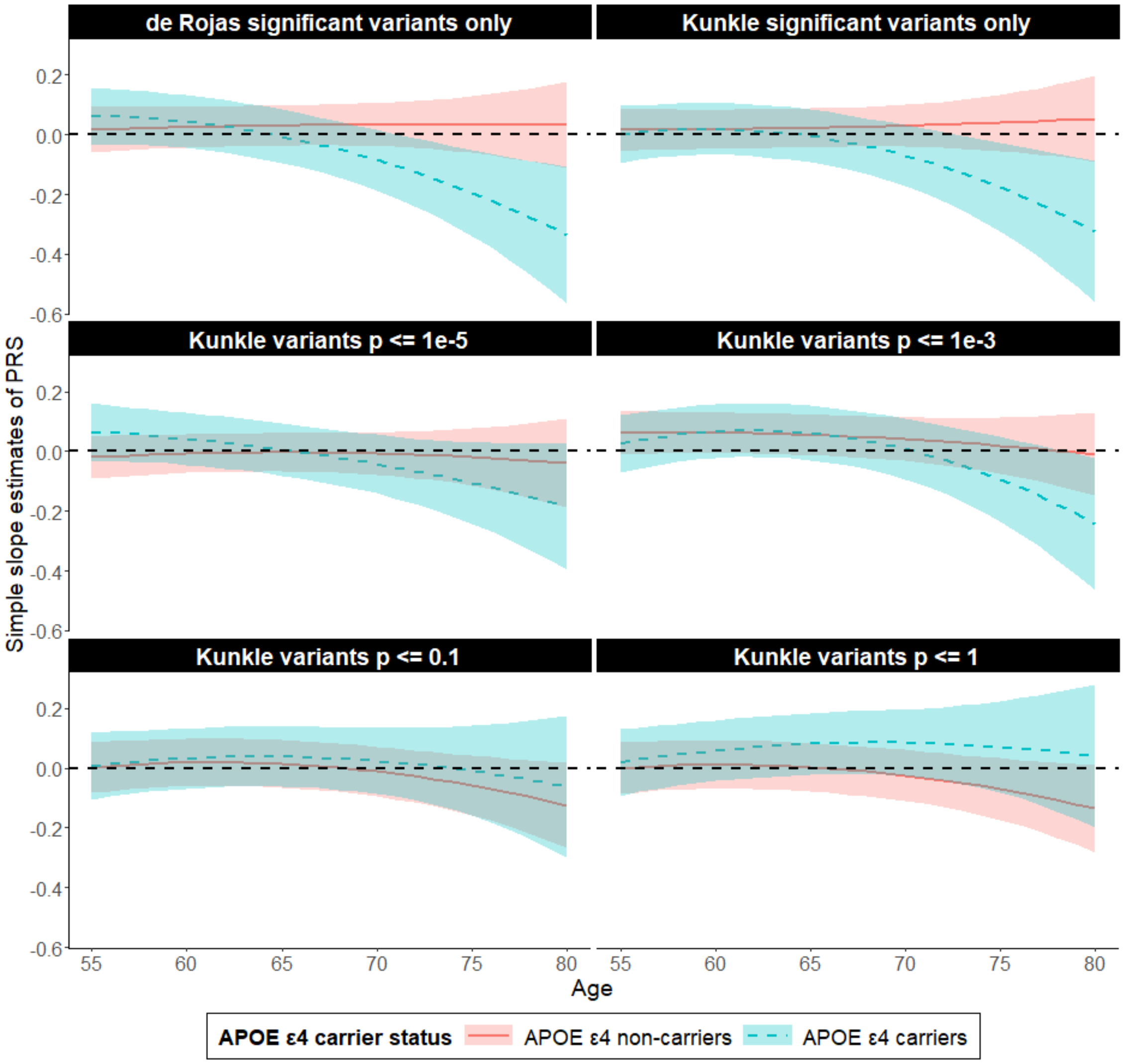


Supplementary figure 1 plotted the simple slope estimates of PRS constructed using different p-threshold for individuals with and without *APOE* ε4 from age 50 to 80 on immediate learning composite score. The red line represents the longitudinal trajectory of simple slope estimates of PRS among *APOE* ε4 non-carriers while the blue line represents simple slope estimates of PRS among individuals with *APOE* ε4. Bands represent 95% confidence intervals. The simple slope estimates are calculated using package "reghelper" in R and were based on the results which were obtained using the linear mixed effect model and adjusted for within-individual/family correlation. In addition to PRS, age (quadratic), *APOE* ε4, and their interactions, additional covariates include gender, education years, practice effect, family history of AD, and the first five principal components. Age is centered at year 65 and education is centered at the mean.

**Supplementary figure 2. Simple slope estimates of PRSs constructed using IGAP GWAS summary statistics with different thresholding parameters on delayed recall cognitive score for individuals with and without *APOE* ε4 and at different age (N = 1,190).**

**Delayed Recall**

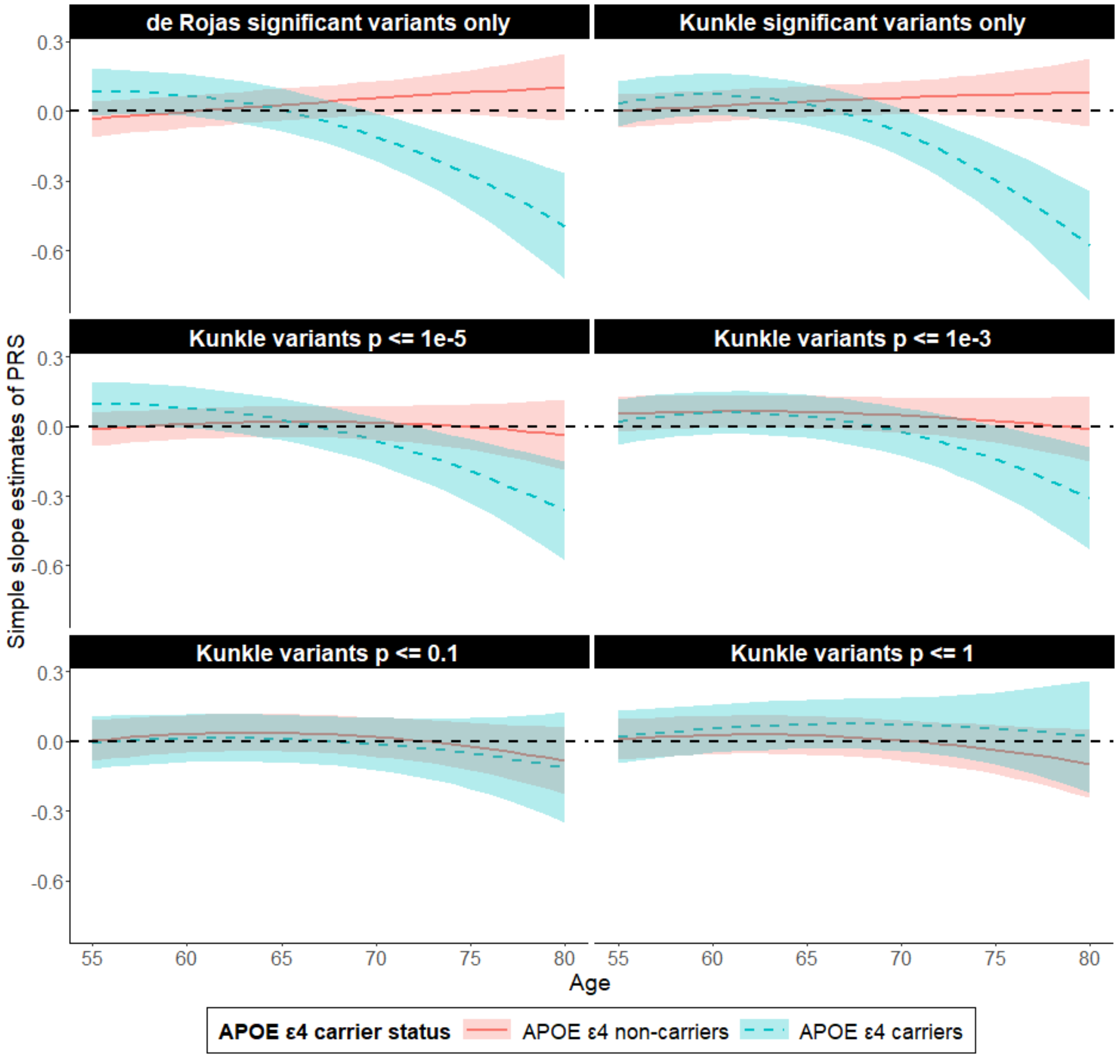


Supplementary figure 2 plotted the simple slope estimates of PRS constructed using different p-threshold for individuals with and without *APOE* ε4 from age 50 to 80 on delayed recall composite score. The red line represents the longitudinal trajectory of simple slope estimates of PRS among *APOE* ε4 non-carriers while the blue line represents simple slope estimates of PRS among individuals with *APOE* ε4. Bands represent 95% confidence intervals. The simple slope estimates are calculated using package "reghelper" in R and were based on the results which were obtained using the linear mixed effect model and adjusted for within-individual/family correlation. In addition to PRS, age (quadratic), *APOE* ε4, and their interactions, additional covariates include gender, education years, practice effect, family history of AD, and the first five principal components. Age is centered at year 65 and education is centered at the mean.

**Supplementary figure 3. Simple slope estimates of PRSs constructed using IGAP GWAS summary statistics with different thresholding parameters on executive function cognitive score for individuals with and without *APOE* ε4 and at different age (N = 1,190).**

#### Executive Function

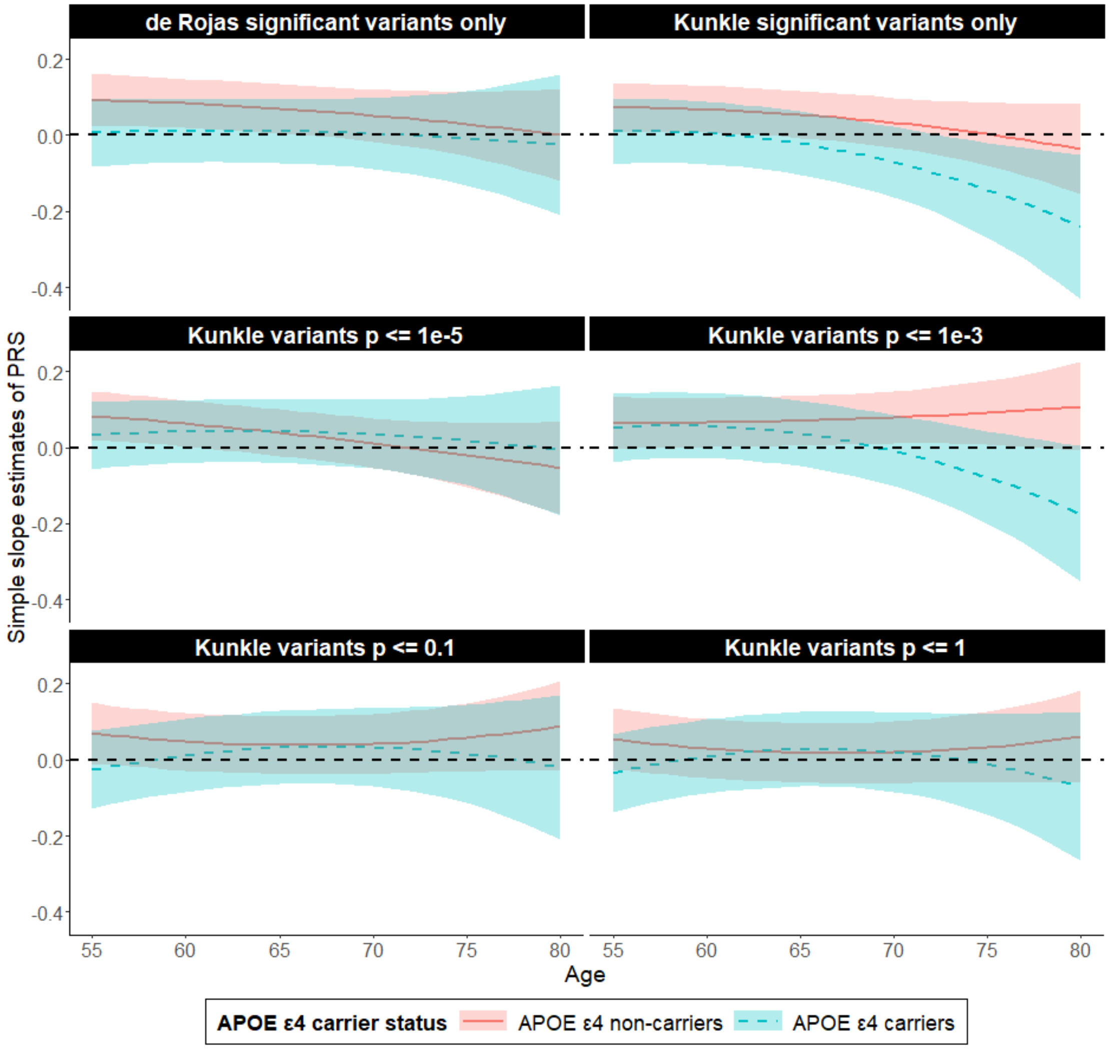


Supplementary figure 3 plotted the simple slope estimates of PRS constructed using different p-threshold for individuals with and without *APOE* ε4 from age 50 to 80 on executive function composite score. The red line represents the longitudinal trajectory of simple slope estimates of PRS among *APOE* ε4 non-carriers while the blue line represents simple slope estimates of PRS among individuals with *APOE* ε4. Bands represent 95% confidence intervals. The simple slope estimates are calculated using package “reghelper” in R and were based on the results which were obtained using the linear mixed effect model and adjusted for within-individual/family correlation. In addition to PRS, age (quadratic), *APOE* ε4, and their interactions, additional covariates include gender, education years, practice effect, family history of AD, and the first five principal components. Age is centered at year 65 and education is centered at the mean.

**Supplementary figure 4. Simple slope estimates of PRSs constructed using IGAP GWAS summary statistics with different thresholding parameters on PACC-3 cognitive score for individuals with and without *APOE* ε4 and at different age (N = 1,190).**

**PACC-3**

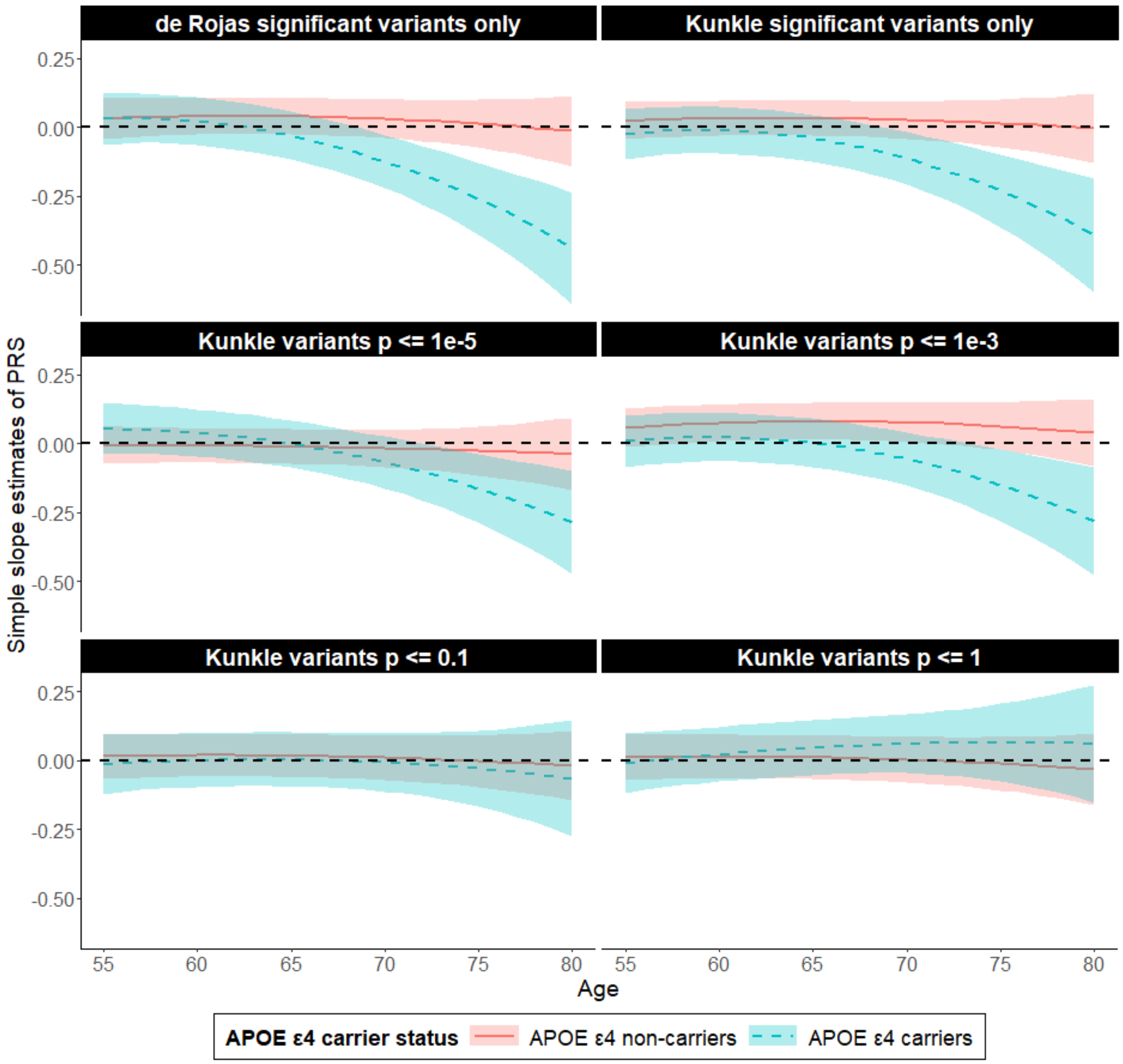


Supplementary figure 4 plotted the simple slope estimates of PRS constructed using different p-threshold for individuals with and without *APOE* ε4 from age 50 to 80 on PACC-3 composite score. The red line represents the longitudinal trajectory of simple slope estimates of PRS among *APOE* ε4 non-carriers while the blue line represents simple slope estimates of PRS among individuals with *APOE* ε4. Bands represent 95% confidence intervals. The simple slope estimates are calculated using package "reghelper" in R and were based on the results which were obtained using the linear mixed effect model and adjusted for within-individual/family correlation. In addition to PRS, age (quadratic), *APOE* ε4, and their interactions, additional covariates include gender, education years, practice effect, family history of AD, and the first five principal components. Age is centered at year 65 and education is centered at the mean. PACC-3 = Preclinical Alzheimer's Cognitive Composite Score-3.

**Supplementary figure 5. Simple slope estimates of $PRS_{Kunkle\_sig}$ on domain specific- and global cognitive score for individuals with different *APOE* genotype and at different age (N = 1,190).**

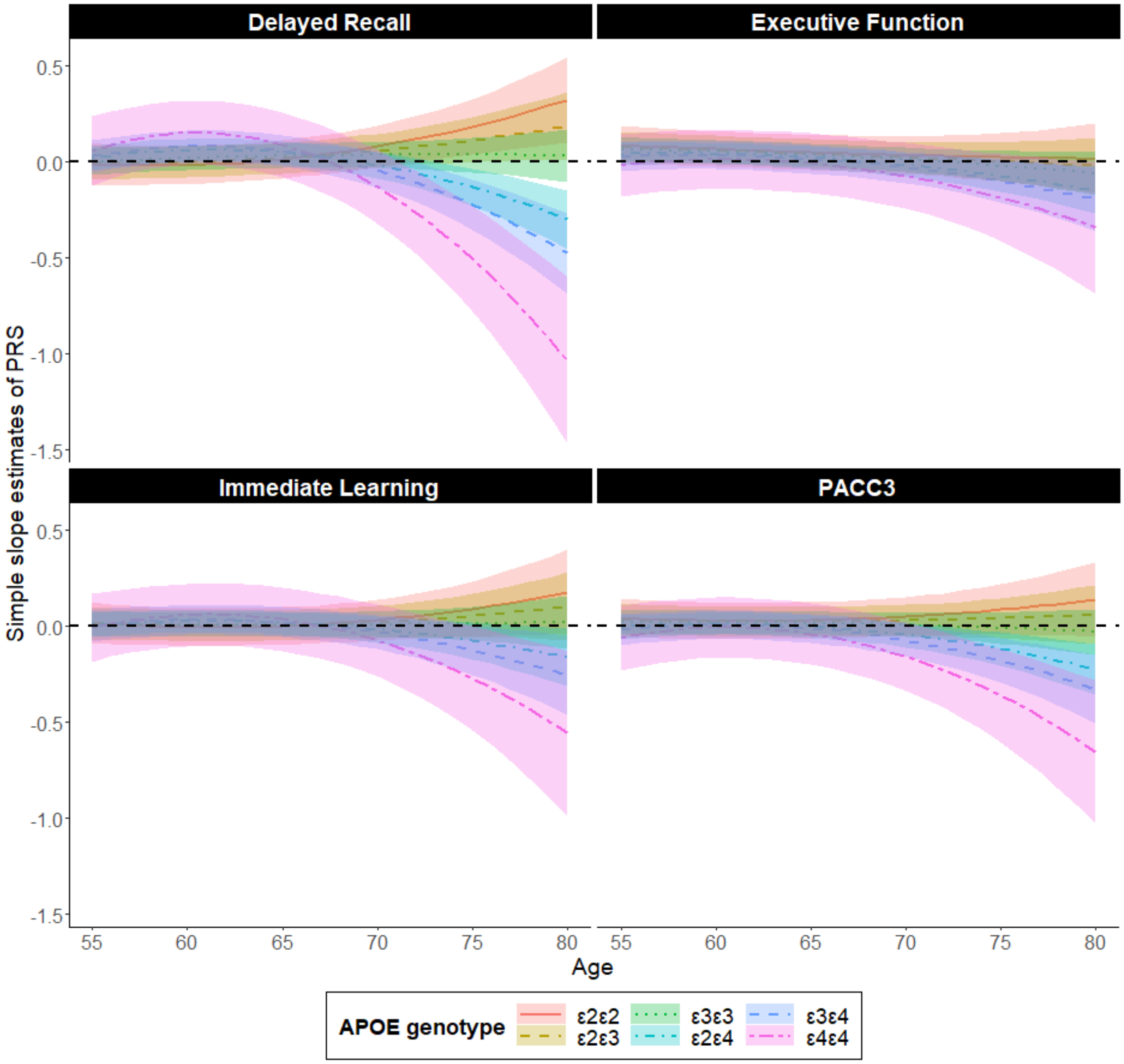


Supplementary figure 5 plotted the simple slope estimates of $PRS_{Kunkle\_sig}$ for individuals with different *APOE* genotypes from age 50 to 80 on global and domain specific cognition score. Each line represents the longitudinal trajectory of simple slope estimates of $PRS_{Kunkle\_sig}$ for individuals with different *APOE* genotypes (as quantified by the *APOE* score). Bands represent 95% confidence intervals. The simple slope estimates are calculated using package "reghelper" in R and were based on the results which were obtained using the linear mixed effect model and adjusted for within-individual/family correlation. In addition to PRS, age (quadratic), *APOE* score, and their interactions, additional covariates include gender, education years, practice effect, family history of AD, and the first five principal components. Age is centered at year 65 and education is centered at the mean. PACC-3 = Preclinical Alzheimer's Cognitive Composite Score-3.

**Supplementary figure 6. Simple slope estimates of $PRS_{Kunkle\_sig}$ on domain specific- and global cognitive score for individuals with and without *APOE* ε4 and at different age in Health and Retirement Study (N = 6,785).**

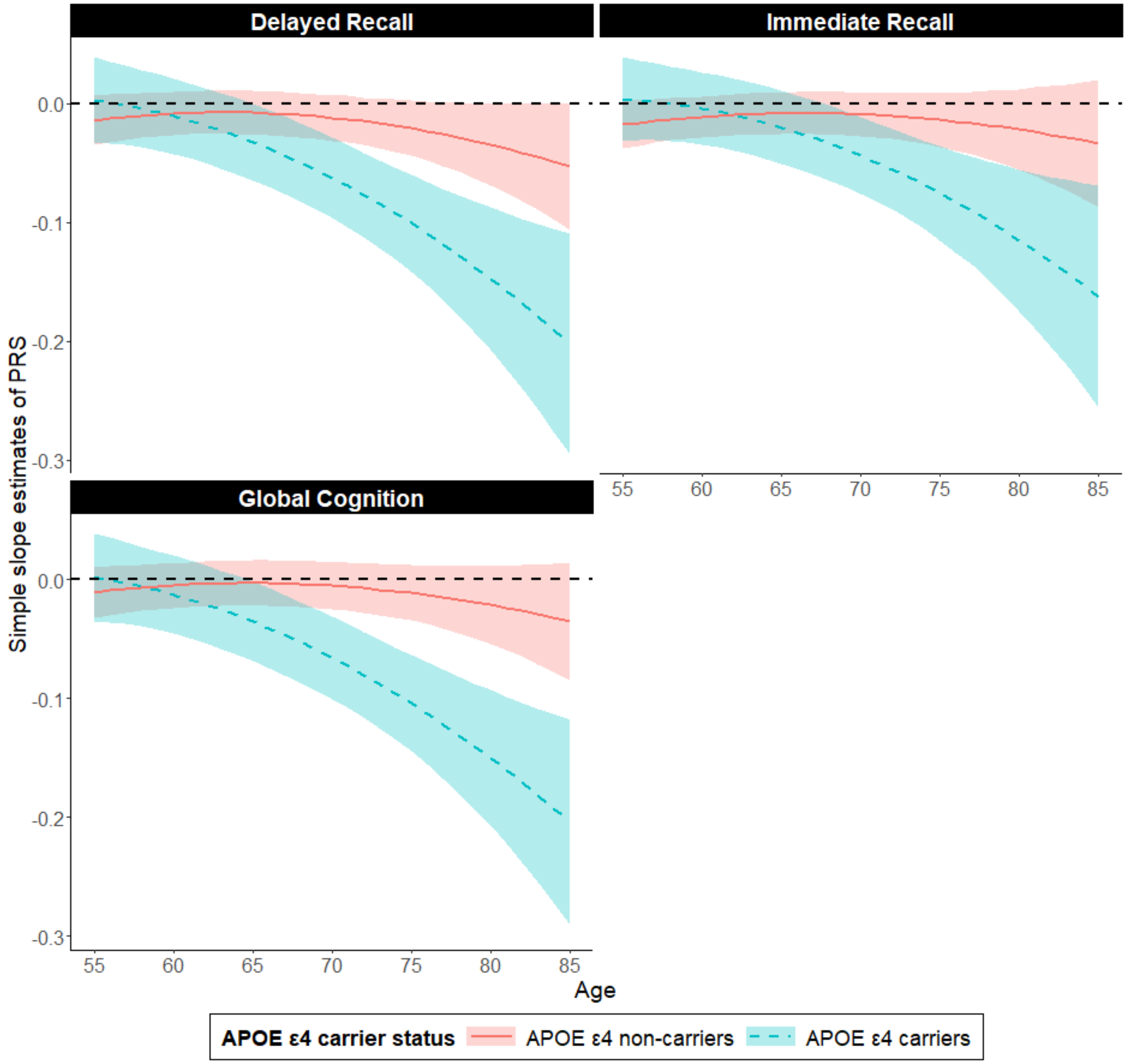


Supplementary figure 6 plotted the simple slope estimates of $PRS_{Kunkle\_sig}$ for individuals with and without *APOE* ε4 from age 50 to 85 on global and domain specific cognition score in the Health and Retirement Study. Inclusion criteria include individuals who were born between 1935 and 1959 (age 40 to 65 at year 2000, not-late babyboomers cohort), non-Hispanic white, whose cognition were not assessed through proxy, and are not classified as “demented” by Langa-Weir Classification of Cognitive Function. The red line represents the longitudinal trajectory of simple slope estimates of $PRS_{Kunkle\_sig}$ among *APOE* ε4 non-carriers while the blue line represents simple slope estimates of $PRS_{Kunkle\_sig}$ among individuals with *APOE* ε4. Bands represent 95% confidence intervals. The simple slope estimates are calculated using package “reghelper” in R and were based on the results which were obtained using the linear mixed effect model and adjusted for within-individual/family correlation. In addition to PRS, age (quadratic), *APOE* ε4, and their interactions, additional covariates include gender, education years, cohort, practice effect, and the first five principal components. Age is centered at year 65 and education is centered at the mean.
